# Early Life Domains as Predictors of Obesity and Hypertension Comorbidity: Findings from the 1970 British Cohort Study (BCS70)

**DOI:** 10.1101/2024.05.13.24307277

**Authors:** S Stannard, RK Owen, A Berrington, N Ziauddeen, SDS Fraser, S Paranjothy, RB Hoyle, N A Alwan

## Abstract

**Background:** Obesity and hypertension are major public health problems and are associated with adverse health outcomes. To model realistic prevention scenarios and inform policy, it may be helpful to conceptualise early lifecourse domains of risk and incorporate such information when predicting comorbidity outcomes. We identify exposures across five pre-hypothesised childhood domains and explore them as predictors of obesity and hypertension comorbidity in adulthood.

**Methods:** The analytical sample included 7858 participants in the 1970 British Cohort Study. The outcome was obesity (BMI of ≥30) and hypertension (blood pressure>140/90mm Hg or self-reported doctor’s diagnosis) comorbidity at age 46. Early life domains included: ‘prenatal, antenatal, neonatal and birth’, ‘developmental attributes and behaviour’, ‘child education and academic ability’, ‘socioeconomic factors’ and ‘parental and family environment’. We conducted prediction analysis of the outcome in three stages:(1) stepwise backward elimination to select variables for inclusion for each domain (2) calculation of predicted risk scores of obesity-hypertension for each cohort member within each domain (3) multivariable logistic regression analysis including domain-specific risk scores, sex and ethnicity to assess how well the outcome could be predicted. We additionally included potential adult predictors of obesity-hypertension comorbidity as sensitivity analysis.

**Results:** Including all domain-specific risk scores in the same model, all five domains were significant predictors of obesity-hypertension comorbidity. The predictive power of the model, measured by the area under the curve (AUC), was 0.63 (95%CI 0.61-0.65). Including adult predictors increase the AUC to 0.68 (95%CI 0.66-0.70), and three early life domains - the parental and family environment domain (OR 1.11 95%CI 1.05-1.17) the socioeconomic factors domain (OR 1.09 95%CI 1.04-1.16), and the education and academic ability domain (OR 1.07 95%CI 1.02-1.13) remained predictors of obesity-hypertension comorbidity.

**Conclusions:** We found three robust domains for predicting obesity-hypertension comorbidity. Interventions that address these early life factors could reduce the burden of comorbidity.

## Introduction

Obesity and hypertension are major public health problems [1–2]. In England, 26% of adults have obesity [3], and 30% of adults have hypertension [4]. Both conditions are associated with morbidities later in the lifecourse, including Type 2 diabetes, heart disease, kidney disease, renal disease, strokes, and some cancers, including breast and bowel cancer [5–8]. There is evidence suggesting that in England and Scotland since 2014, obesity and excess body fat have contributed to more deaths among people in middle- and old-age than smoking [9]. On a global level, in 2019 and across 204 countries, the leading Level 2 risk factor for attributable deaths was high systolic blood pressure, and between 2010 to 2019, one of the largest increases in risk exposure was for high body-mass index [10].

Obesity and hypertension are closely related and often co-occur [11], for example each condition occurs at higher frequency with the other than in a population free of either [12]. Research indicates that obesity accounts for 60-70% of hypertension, and individuals with obesity are 3.5 times more likely to have hypertension compared to normal weight individuals [13,14]. The combination of both conditions significantly increases the likelihood of adverse health outcomes such as cardiovascular disease, reduced sexual function, quality of life and mortality [12,15,16]. Previous literature has also identified obesity and hypertension as common sentinel conditions, defined as the first long-term condition in the development of multiple long-term conditions [17–20].

A substantial body of evidence suggests that experiences in early life are crucial in determining outcomes such as obesity and hypertension. The aetiology of chronic disease has been strongly linked with environmental exposures in utero and early life [21]. Socioeconomic disadvantage in early life is also strongly related to obesity and hypertension [22]. Analyses of the Hertfordshire cohort study demonstrated that paternal social class was associated with future multimorbidity, including hypertension [23]. In the Aberdeen Children of the 1950s cohort, lower father’s social class at birth was associated with early-onset multimorbidity, including hypertension [24], and in the 1970 British Cohort Study those with fathers from unskilled occupational groups (vs. professional) at birth had 43% higher risk of early-onset (age 46-48) multimorbidity including hypertension [25].

Many wider determinants acting in childhood are likely to increase the risk of disease in adulthood, and in previous research [26,27] we conceptualised exposures across 12 pre-defined childhood domains covering a range of social, economic, developmental, educational and environmental factors. Most previous research focuses on single exposure-outcome relationships, potentially to reduce statistical complexity, or to focus policy attention onto a specific aspect. However, focussing on single exposures does not reflect the reality and complexity of the early life course given children are likely to be exposed to combinations of intersectional factors across these domains, often simultaneously. We argue that analyses must begin to explore exposures as domains (i.e., a group of variables that represent an overarching theme) rather than the individual variables that form its components for three reasons. First, to provide a combined exposure measure that reflects multiple variables in the data rather than performing multiple statistical testing using all of the components in relation to the study outcomes. Second, to conceptualise the components within wider early life domains provides a better reflection of the childhood conditions in which people grow up. Third considering domains will better inform interventions and policy in childhood as incorporating information from multiple early life domains into the same analysis may help us understand the combined effects of different experiences across a range of early life domains on developing long-term conditions. This can provide actionable insights into developing complex multi-domain interventions in childhood that may help support people to live more healthily for longer across the lifecourse.

In this paper we look to address two research questions. Firstly, which early life domain(s) contribute to predicting the risk of obesity-hypertension comorbidity at mid-life? Secondly, which early life domain(s) are more important than others in explaining the variability in obesity-hypertension occurrence?

To achieve this, we aimed to predict the outcome of obesity-hypertension comorbidity using five early lifecourse domains. In order to do this while weighting the components of each domain, we also aimed to produce -as a first stage of the analysis-predicted risk scores of the outcome for each of the five pre-defined early life domains [26,27]. In a sensitivity analysis we included potential adult risk factors of obesity-hypertension comorbidity (number of days of exercise per week, highest educational qualification, weekly income, number of cigarettes smoked daily, hours spent on a weekday watching television, hours on a weekday spent on the internet (not for work related reasons), cohabitating with a partner, alcohol consumption and use of e-cigarettes) to explore which early life domains matter most for obesity-hypertension comorbidity taking account of such factors.

This work forms part of a larger aim to model targeted multimorbidity prevention scenarios as part of the Multidisciplinary Ecosystem to study Lifecourse Determinants and Prevention of Early-onset Burdensome Multimorbidity (MELD-B) project [28].

## Methods

### Dataset

We used the 1970 British Cohort Study (BCS70) [29] that has followed 17196 cohort members in England, Scotland, Wales born in one week in 1970; to date, there have been 10 sweeps of data collection – 4 in childhood and 6 in adulthood. The comorbidity outcome of obesity and hypertension was measured at age 46 within a biomedical sweep with measurements conducted by a research nurse. All other variables were collected either at birth or age 10.

### Outcome

The outcome was a combined obesity-hypertension phenotype at age 46. Blood pressure was measured via three systolic and diastolic blood pressure readings during a single appointment and administered by a research nurse. Hypertension was defined as an average blood pressure reading of over 140/90 mm Hg. We additionally classified hypertension if a participant reported (at age 46) that they had received a doctor’s diagnosis of high blood pressure or hypertension, even if the blood pressure measurement was less than 140/90 mm Hg, since diagnosed hypertension may be accompanied by intake of antihypertensive medication, thus lowering blood pressure readings at the time of cohort measurement. Body mass index (BMI) was calculated via height and weight measurements taken during the same nurse appointment using the following formula: BMI = weight (kg) / height (m)^2^. Obesity was defined as a BMI of 30 or over. The obesity-hypertension comorbidity variable was considered as a binary (no/yes) variable.

### Exposures (Five pre-hypothesised domains)

We previously conceptualised 12 domains of early life risk factors of future multimorbidity risk informed through a scoping literature/policy review and patient and public engagement [26]. In this paper we focus our analysis on 5 out of the 12 domains chosen because they showed unadjusted associations with the outcomes:

1. *Prenatal, antenatal, neonatal and birth domain* focused on the period from conception to the onset of labour, the circumstances and outcomes surrounding a birth, and the period immediately following birth.
2. *Developmental attributes and behaviour domain* focused on the developmental markers of children relating to cognition, coordination, personality types and behavioural traits.
3. *Child education and academic ability domain* related to the process of learning and educational achievement, especially in educational settings, and the knowledge an individual gains from these educational institutions.
4. *Socioeconomic factors domain* included factors relating to differences between individuals or groups of peoples caused mainly by their social and economic situation.
5. *Parental and family environment domain* incorporated the interactions between children and care givers, parenting styles, parental beliefs, attitudes and discipline, and wider family factors such as kin networks.

Supplementary Materials Table 1 includes all the variables that were initially considered for each domain. These variables were selected and categorised based on a previous data audit and PCA analysis [27] that identified early-life variables from multiple sweeps of data that fitted into five early-life domains of future multimorbidity risk. This previous work reduced the dimensionality of the data and structured each of the five domains into mutually exclusive groups of variables based on similar characteristics [27].

**Table 1.**
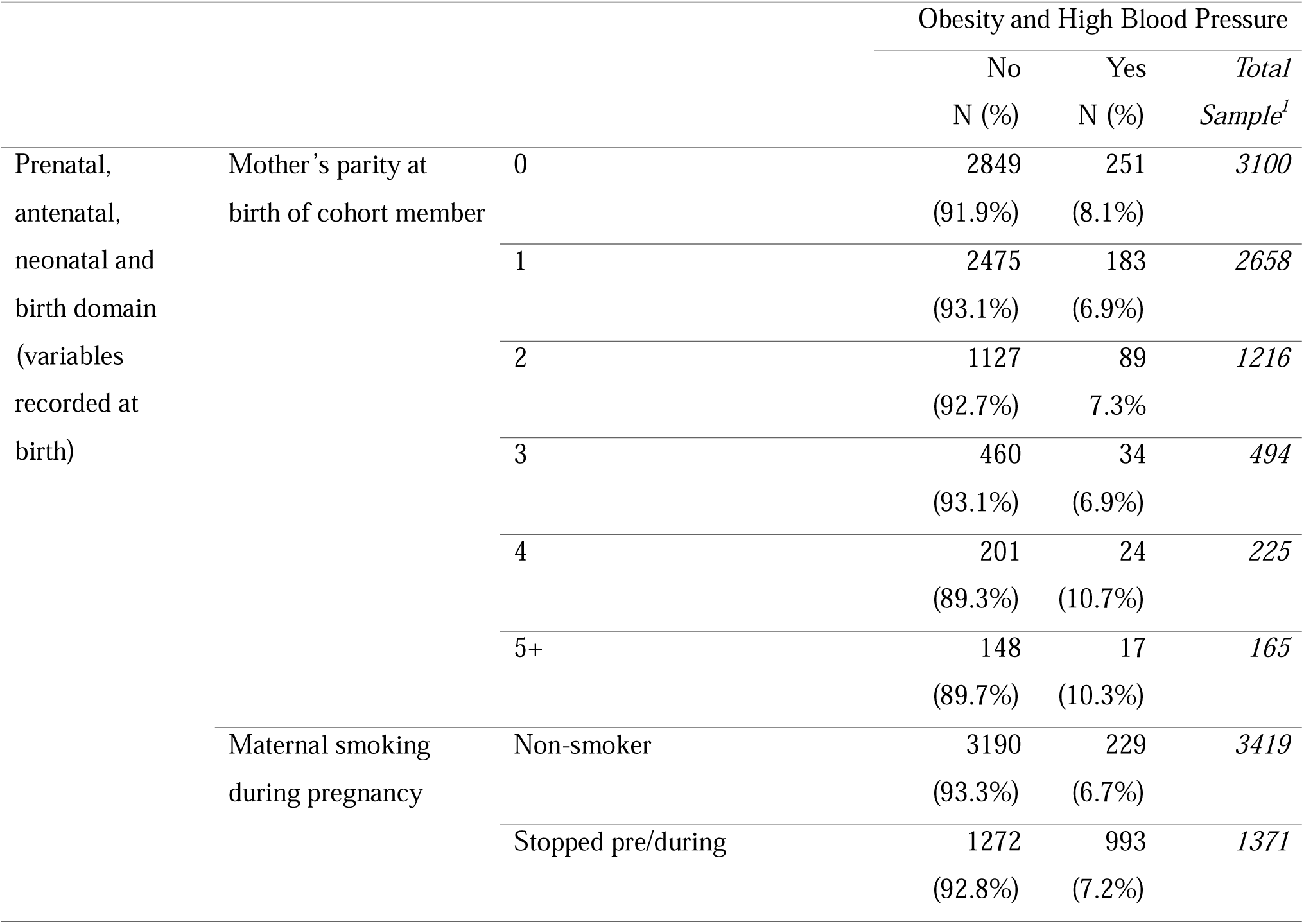
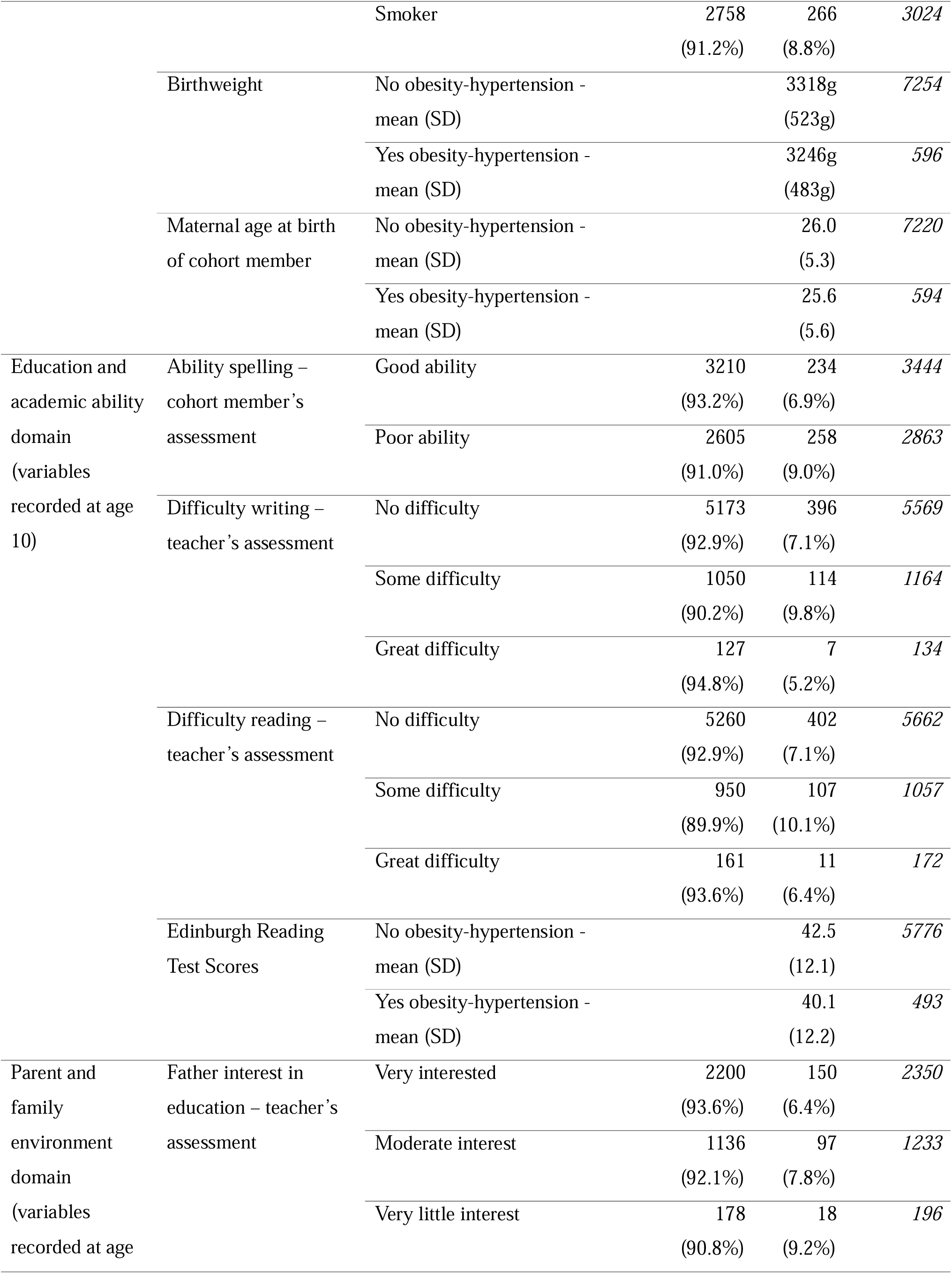
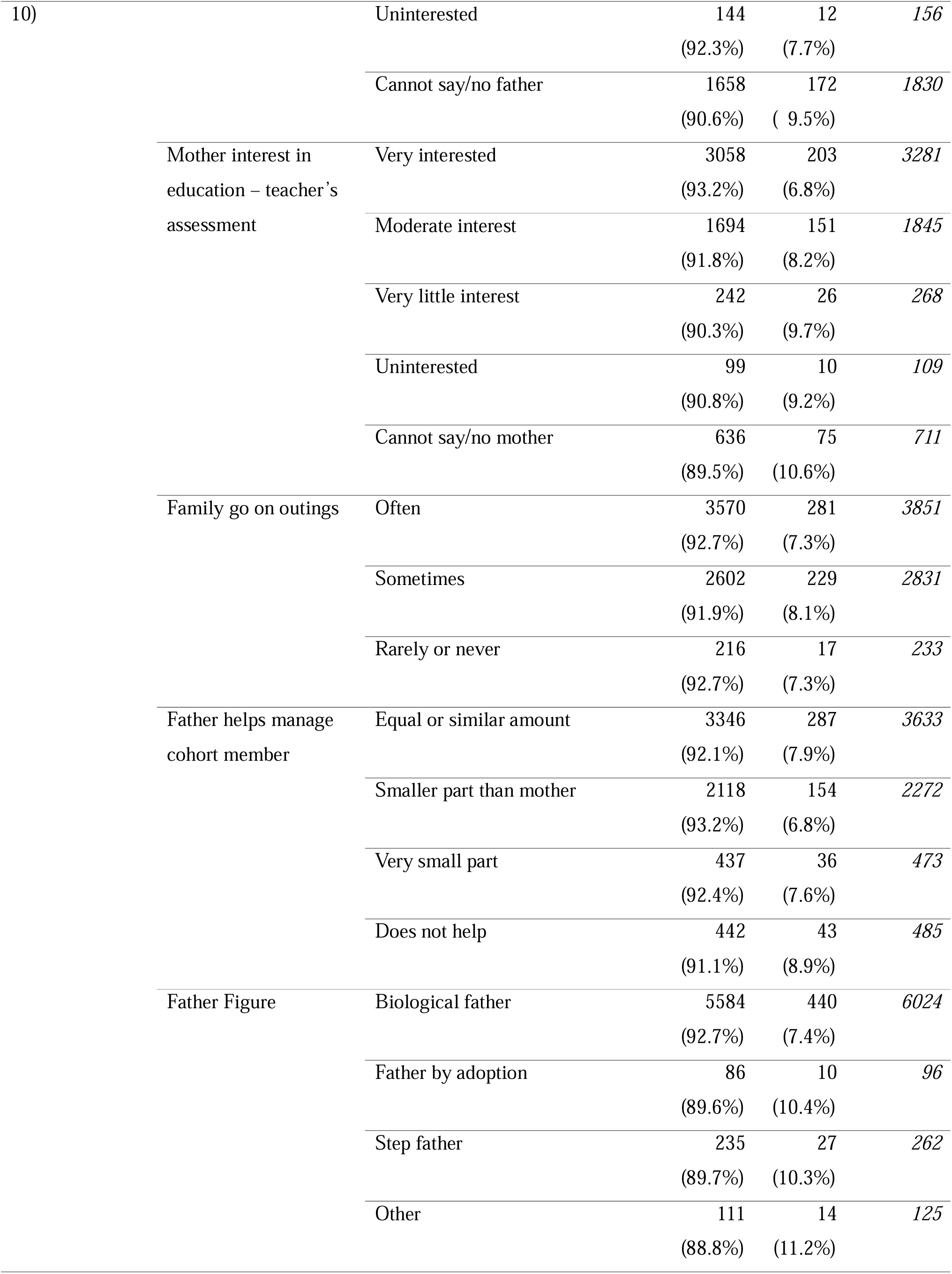
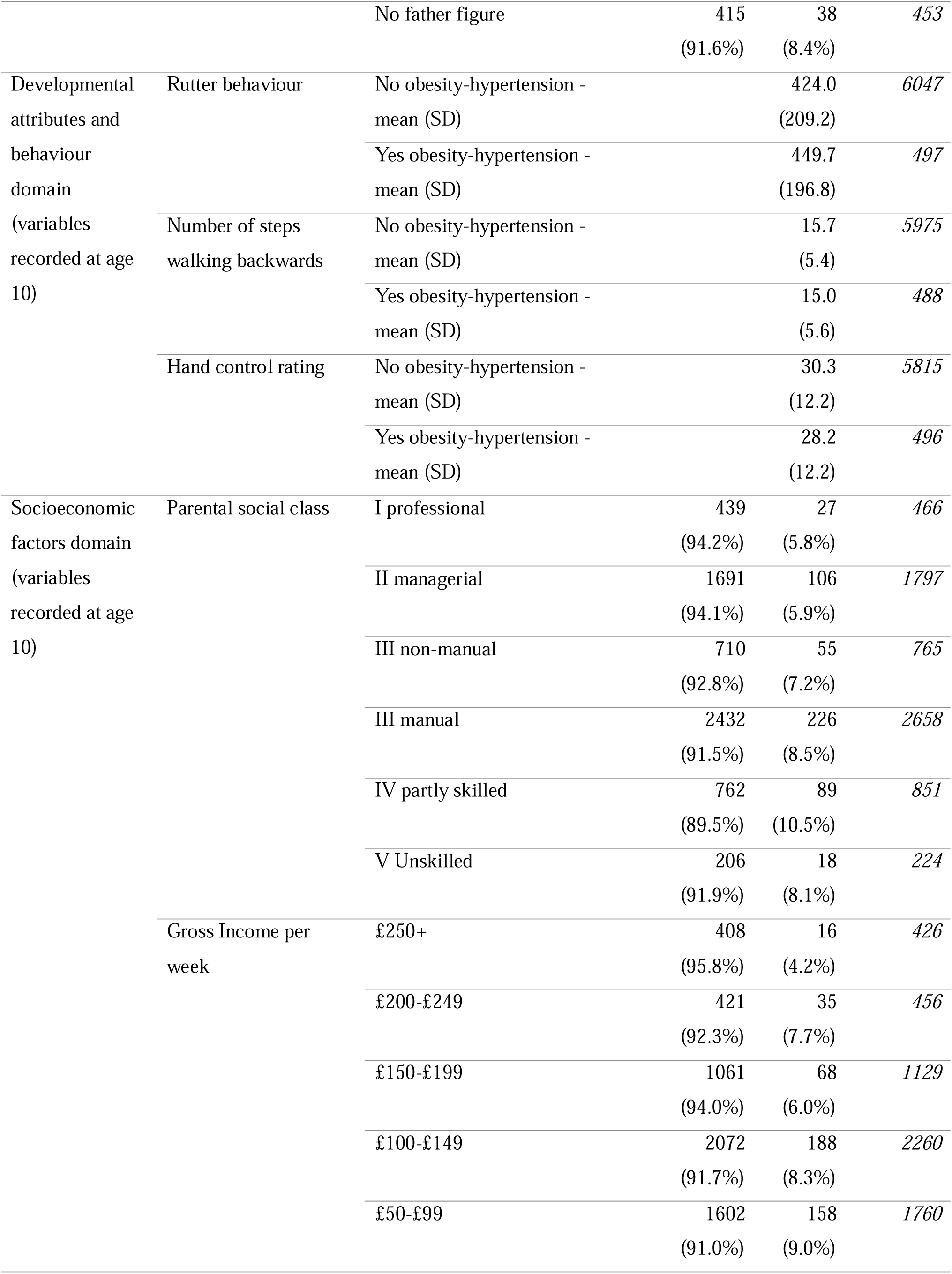
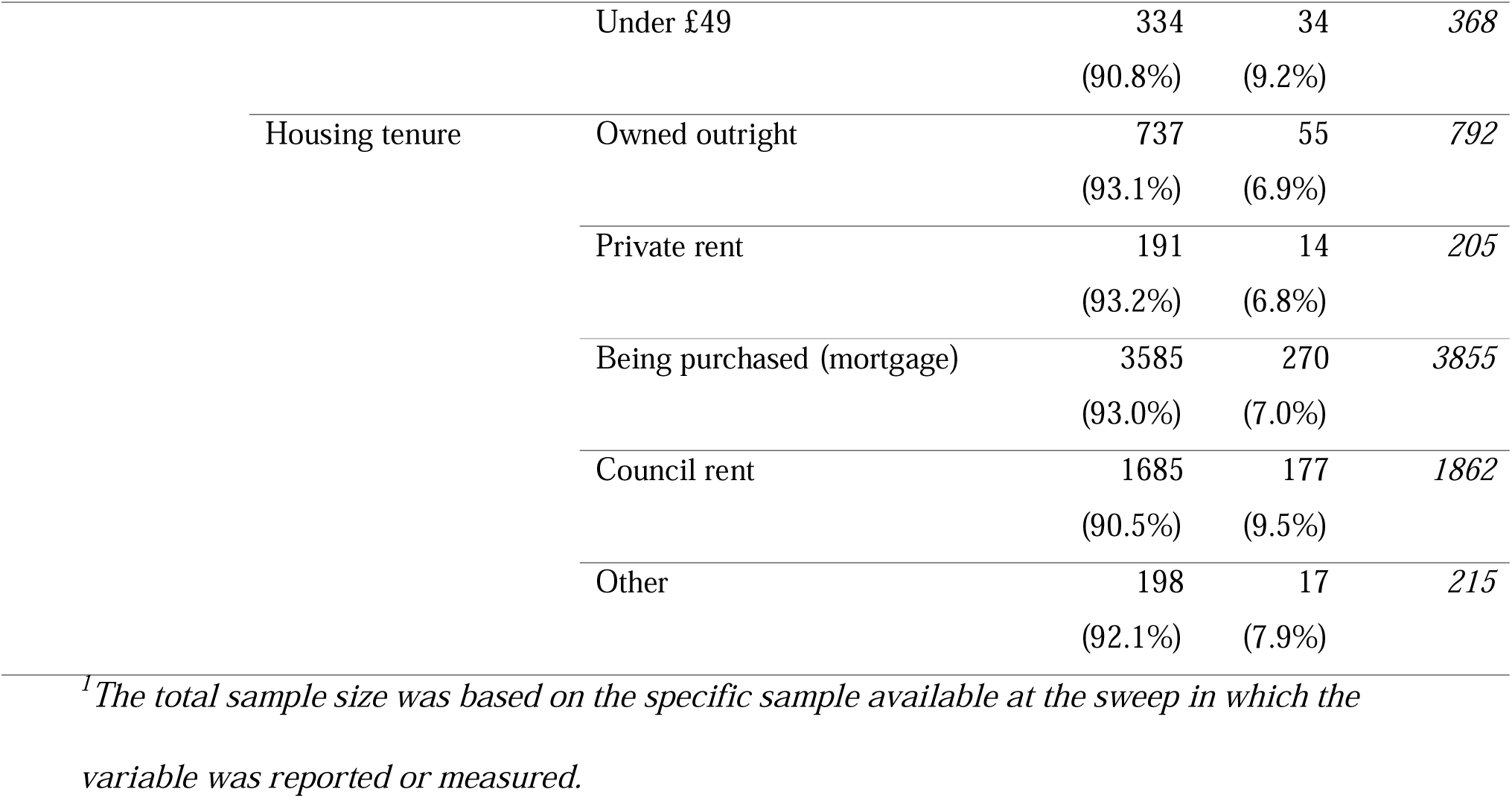
Step 1: Variables retained in the five domains risk scores following stepwise backwards elimination, and the prevalence of obesity-hypertension comorbidity at age 46.

### Analytical sample

The analytical sample included all cohort members who had measured BMI and blood pressure at age 46 (n = 7858); this represented 45.7% of the original birth cohort. To preserve sample size and reduce bias in the estimates due to missing data we used multiple imputation. Multiple imputation was conducted by chained equations for missing observations at birth, age 10, and 46 [30]. 50 imputation cycles were constructed under the missing-at-random assumption [31–33], which has been found to be highly plausible in the British birth cohorts [34]. All variables were included in the imputation process. The outcome was included in the imputed models, but imputed outcome values were not used. For reference we include results based on complete case analysis in Supplementary Materials Tables 2 and 9.

**Table 2.**
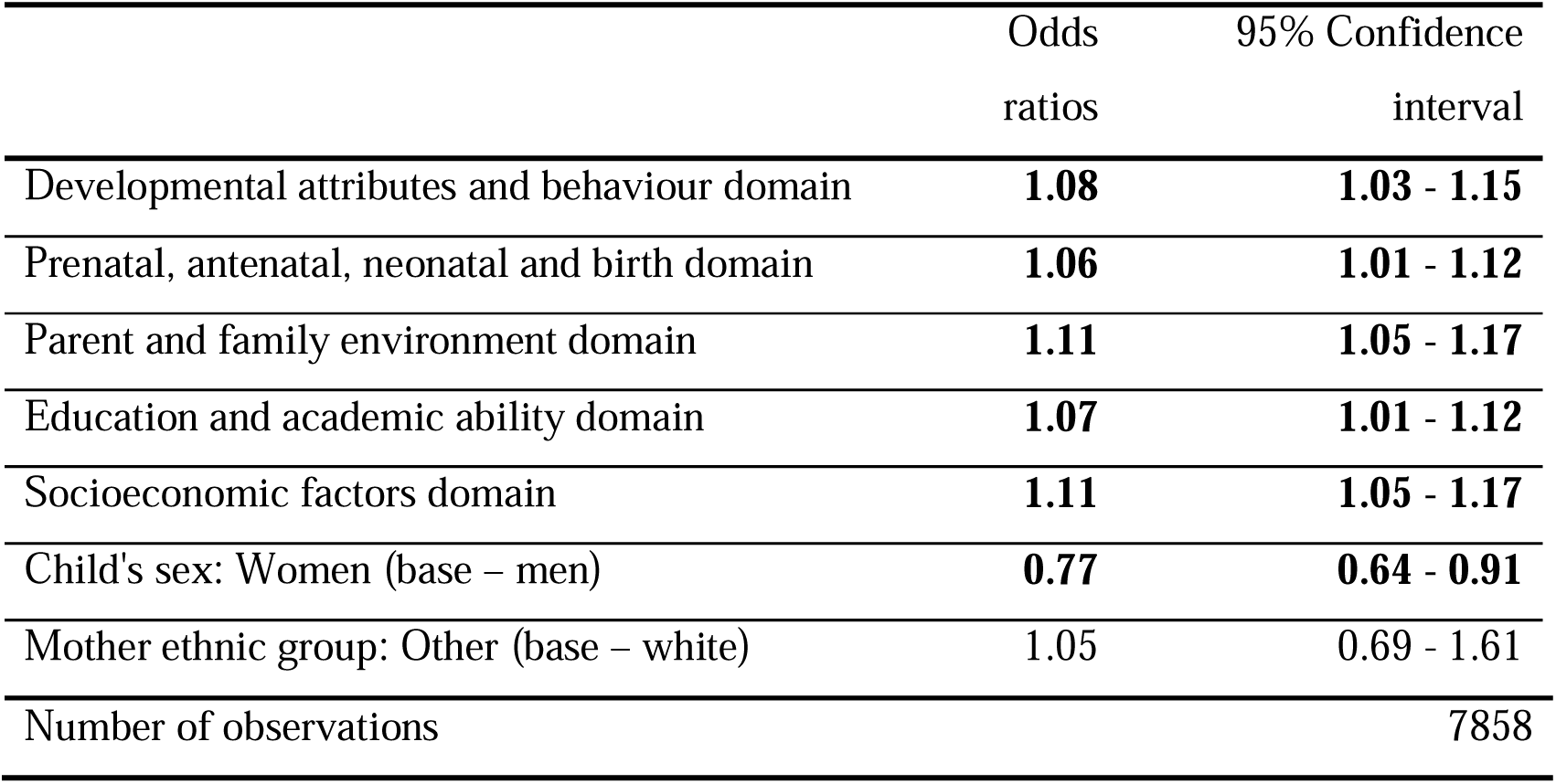
Step 3: Odds ratios of obesity-hypertension at age 46 in relation to domain-specific risk score of obesity-hypertension for five early life domains. Multiple imputed data (50 Imputations)

#### Statistical analysis

##### Step 1: Stepwise backwards elimination to select variables for inclusion separately for each domain

Firstly, stepwise backward elimination conducted on multiple imputed data, was used to select variables for inclusion separately for each domain. This method started with all potential variables identified for each domain as outlined in Supplementary Materials Table 1, then variables were removed sequentially based on a series of hypothesis tests. In backward elimination, variables were removed sequentially if the p-value for a variable exceeded the specified significance level which was set at 0.157. This level was chosen conservatively to reduce the risk of overfitting and is the equivalent to the Akaike information criterion (AIC) [35,36]. Table 1 identifies the retained variables following stepwise backwards elimination for each domain in relation to the outcome of obesity-hypertension comorbidity at age 46, the descriptive statistics were based on the specific sample available at the sweep in which the variable was reported or measured.

##### Step 2: Predicted risk scores of obesity-hypertension comorbidity for each cohort member within each domain

Secondly, logistic regression models then explored the relationship between retained variables within each domain following stepwise backwards elimination and odds of obesity-hypertension comorbidity. Based on this logistic regression modelling, and using the ‘predict’ function in STATA [37], predicted risk scores of the obesity-hypertension comorbidity outcome for each cohort member within each domain were calculated. In other words, each cohort member has five predicted score values in relation to the obesity-hypertension comorbidity outcome, one for each of the five domains. These predicted risk scores for each individual, and within each domain, were centred on the mean predicted risk score within that domain, and were bound between −1 and 1. A Pearson correlation matrix then explored the correlation between domain-specific risk scores. We generated predicted risk scores of obesity-hypertension for each of the five early life domains to account for the weight of each domain component in relation to the outcome rather than assuming each component contributes an equal weight to the outcome.

##### Step 3: A prediction model including five domain-specific risk scores

The third step involved focussing on how well obesity-hypertension could be predicted including all five domain-specific risk scores produced in step 2 using a multivariable logistic regression model with the ‘predict function’, and adding sex (at birth) and ethnicity. We produced the area under curve (AUC) statistic to assess the predictive performance of this model. Odds ratios and confidence intervals of the five domains within this model were used to identify the strongest domains that acted as predictors for obesity and hypertension comorbidity taking into account the effect of the other domains.

##### Step 4: A prediction model including five domain-specific risk scores and adult factors (sensitivity analyses)

The fourth step involved performing step 3 with the inclusion of adult factors that are potentially linked to both the exposures and the outcome. These were recorded at age 46 including number of days of exercise per week, highest educational qualification, occupational social class, weekly income, number of cigarettes smoked daily, hours spent on a weekday watching television, hours on a weekday spent on the internet (not for work related reasons), cohabitating with a partner, alcohol consumption and use of e-cigarettes.

#### Ethical considerations

Ethics approval for the MELD-B project has been obtained from the University of Southampton Faculty of Medicine Ethics committee (ERGO II Reference 66810).

## Results

### Step 1: Stepwise backwards elimination to select variables for inclusion in each domain

Among the 7858 cohort members at age 46, 597 (7.6%) had obesity-hypertension comorbidity at age 46. Table 1 identifies the retained variables following stepwise backwards elimination for each domain in relation to the outcome of obesity-hypertension comorbidity at age 46. As shown in Table 1, in the socioeconomic factors domain, 9.2% of cohort members whose gross family income at age 10 (per week) was below £49 had obesity-hypertension at age 46, compared to 4.2% of cohort members whose gross family income was above £250. In the prenatal, antenatal, neonatal domain, 8.8% of cohort members whose mothers smoked during pregnancy had obesity and hypertension at age 46 compared to a 6.7% prevalence in non-smokers. In the education and academic ability domain 10.1% of cohort members who reported some difficulty with reading at age 10 had obesity-hypertension at age 46 compared to a prevalence of 7.1% in those with no difficulty. Finally, for the parent and family environment domain of cohort members with a father by adoption (10.4%), stepfather (10.3%) or another father figure such as a grandparent (11.2%) at age 10 had obesity-hypertension at age 46 compared to 7.4% in those living with a biological father.

### Step 2: Generation of domain-specific predicted risk scores of obesity-hypertension comorbidity for each cohort member

Supplementary Materials Figure 1. presents a histogram of the distribution of domain-specific predicted risk scores, generated in step 2, following logistic regression modelling that explored the relationship between retained variables following stepwise backwards elimination (step 1) and obesity-hypertension comorbidity for the five early life domains. In Supplementary Materials Tables 4-8, we include the regression coefficients of obesity-hypertension for these models, and for each domain separately. The domain risk scores were centred on the mean predicted risk score (for each domain) and bound between −1 and 1. As shown the largest range of domain predicted risk scores was for the prenatal, antenatal, neonatal and birth domain (range −0.05 – 0.14) and the smallest range was for the parental and environmental domain risk score (range −0.03 – 0.08).

Given there was likely to be correlation between domain risk scores, in Supplementary Materials Table 3 we present a Pearson correlation matrix exploring correlation across domains. The highest correlation was between both the developmental attributes and behaviour domain and the child education and academic ability domain (coefficient 0.29), and between the parental and family environment domain and the socioeconomic factors domain (coefficient 0.32), and therefore the correlation between predicted risk scores for each domain was low.

**Table 3.** Step 3: Odds ratios of obesity-hypertension at age 46 in relation to domain-specific risk score of obesity-hypertension for five early life domains including adult predictors.

|  | OR | 95% Confidence<br>Interval |
| --- | --- | --- |
| Developmental attributes and behaviour domain | 1.07 | 1.00 - 1.13 |
| Prenatal, antenatal, neonatal and birth domain | 1.05 | 0.99 - 1.11 |
| Parent and family environment domain | **1.11** | **1.05 - 1.17** |
| Education and academic ability domain | **1.07** | **1.02 - 1.13** |
| Socioeconomic factors domain | **1.09** | **1.04 - 1.16** |
| Child's sex: Women (base – men) | **0.75** | **0.62 - 0.91** |
| Mother ethnic group: Other (base – white) | 0.89 | 0.58 - 1.37 |
| Number of cigarettes smoked a day | 0.99 | 0.98 - 1.01 |
| TV hours per weekday: 1 to 2 hours (base - 1 hour or less) | **1.72** | **1.24 - 2.39** |
| 2 to 3 hours | **1.46** | **1.03 - 2.06** |
| 3 to 4 hours | **1.49** | **1.02 - 2.17** |
| More than 4 hours | **1.90** | **1.26 - 2.86** |
| Internet hours per weekday: 1 to 2 hours (base -1 hour or less) | 1.12 | 0.91 - 1.38 |
| 2 to 3 hours | **1.37** | **1.04 - 1.80** |
| 3 to 4 hours | 1.37 | 0.95 - 1.98 |
| More than 4 hours | **1.47** | **1.03 - 2.09** |
| Cohabiting: No (base – yes) | 1.04 | 0.85 - 1.27 |
| Alcohol consumption: Does not drink alcohol (base – normal) | **1.39** | **1.08 - 1.81** |
| High risk drinking | **1.23** | **1.00 - 1.51** |
| Social class: Lower managerial (base - higher managerial) | 1.10 | 0.82 - 1.49 |
| Intermediate occupations | 1.12 | 0.77 - 1.63 |
| Small employer | 0.95 | 0.63 - 1.43 |
| Lower supervisor | 1.04 | 0.71 - 1.55 |
| Semi-routine | 0.89 | 0.59 - 1.35 |
| Routine | 0.82 | 0.52 - 1.28 |
| Not stated | 1.15 | 0.80 - 1.67 |
| Not applicable | 0.75 | 0.48 - 1.17 |
| Days exercise per week: 1 (base – 0) | 0.75 | 0.54 - 1.05 |
| 2 | **0.74** | **0.55 - 0.99** |
| 3 | **0.74** | **0.55 - 0.99** |
| 4 | **0.67** | **0.47 - 0.97** |
| 5 | 0.80 | 0.59 - 1.07 |
| 6 | **0.56** | **0.32 - 0.98** |
| 7 | **0.54** | **0.41 - 0.72** |
| Educational qualification: GCSE (base - no qualification) | 1.32 | **1.06 - 1.64** |
| A/AS level | 1.24 | 0.81 - 1.91 |
| Diploma | 1.39 | 0.99 - 1.95 |
| Degree | 1.20 | 0.89 - 1.63 |
| Higher degree | 1.01 | 0.58 - 1.75 |
| Weekly income | 1.00 | 0.99 - 1.00 |
| Managing financially: doing all right (base - doing well) | 1.06 | 0.86 - 1.30 |
| Just about getting by | 1.24 | 0.95 - 1.61 |
| Finding it quite difficult | **1.72** | **1.16 - 2.53** |
| Finding it very difficult | 1.65 | 0.99 - 2.78 |
| E-cigarettes: Previously smoked (base- never smoked) | 0.97 | 0.62 - 1.39 |
| Occasionally smoke | 1.43 | 0.97 - 2.14 |
| Smoke daily | 0.82 | 0.56 - 1.21 |
| Number of observations |  | 7858 |

### Step 3: Prediction modelling of obesity-hypertension including five domain-specific risk scores

We calculated the area under curve (AUC) (included in Supplementary Materials 10) for the prediction model that included all five domain specific risk scores (produced in step 2), ethnicity and sex. Overall, the area under the curve was 0.63 (95%CI 0.61-0.65). Table 2 presents the odds ratios of obesity-hypertension at age 46 for the prediction model that included all five domain specific risk scores (produced in step 2), sex and ethnicity. As shown, all domain risk scores were significant predictors of obesity-hypertension comorbidity, with the strongest being the parental and family environment domain (OR 1.11 95%CI 1.05-1.17) and the socioeconomic factors domain (OR 1.11 95%CI 1.05-1.17). The weakest domain predictors included the prenatal, antenatal, neonatal and birth domain (OR 1.06 95%CI 1.01-1.12) and the education and academic ability domain (OR 1.07 95%CI 1.01-1.12).

### Step 4: Prediction modelling of obesity-hypertension comorbidity including five domain-specific risk scores and potential adult predictors (sensitivity analysis)

Table 3 presents the odds ratios of obesity-hypertension at age 46 for the prediction model that included all five domain specific risk scores (produced in step 2) ethnicity, sex and adult predictors. Overall the inclusion of adult predictors increased the area under the curve to 0.68 (95%CI 0.66-0.70) (Supplementary table 11). As shown, after the inclusion of adult predictors, the prenatal, antenatal, neonatal and birth domain (OR 1.05 95%CI 0.99-1.11) and the developmental attributes and behaviour domains (OR 1.07 95%CI 1.00-1.13) were no longer significant predictors of obesity-hypertension comorbidity. The other three domains remained significant predictors for obesity and hypertension comorbidity. These domains included the parental and family environment domain (OR 1.11 95%CI 1.05-1.17), the socioeconomic factors domain (OR 1.09 95%CI 1.04-1.16), and the education and academic abilities domain (OR 1.07 95%CI 1.02-1.13).

## Discussion

Using the BCS70 data, early life domains showed associations with obesity-hypertension comorbidity at age 46 even after accounting for each other, with the strongest domain predictors for obesity-hypertension being the parental and family environment domain and the socioeconomic factors domain. After accounting for adult predictors including number of days of exercise per week, highest educational qualification, occupational social class, weekly income, number of cigarettes smoked daily, hours spent on a weekday watching television, hours on a weekday spent on the internet (not for work related reasons), cohabitating with a partner, alcohol consumption and use of e-cigarettes, associations with the outcome for the parental and family environment domain, the socioeconomic factors domain, and the education and academic abilities domain remained robust.

In this paper we have quantified the effect of a combination of risk factors in early life in the shape of life domains on comorbidity outcomes. We recognise that prevention policy choices are rarely feasible or practical for one isolated risk factor and thinking in terms of domains may help addressing the early wider determinants of later health. Therefore, our motivation for this analysis stems from the observation that health research tends to investigate single exposure-outcome relationships, while children’s experiences across a variety of early lifecourse behavioural, social, economic and transgenerational domains are intersecting, and therefore research needs to be able to incorporate information from multiple domains into the same analysis. However, it is also important that health research reflects on modifiable risk factors for ill health that focus on both direct and indirect factors including wider systemic and structural determinants of disease and health inequalities [38].

Previous research has indicated that early life factors, including environmental exposures in utero, socioeconomic disadvantage, and father’s occupation are associated to the individual outcomes considered in this paper [21–25]. We extended this previous work to demonstrate that combinations of early-life risk factors represented through domains are also associated to obesity-hypertension comorbidity. In particular our results support research for different cohort studies including the Hertfordshire cohort study and the Aberdeen Children of the 1950s cohort suggesting that early life socioeconomic disadvantage including paternal social class and occupational social class are key in shaping multimorbidity [22–25]. We have demonstrated that this relationship remains even after considering the role of other early life domains and adult predictors.

The finding that the parental and family environment domain remains important indicate that there may be lifelong health impacts stemming from the interaction between children and the primary care giver, parenting styles, parental beliefs, attitudes and discipline, and wider family factors such as kin networks in childhood.

Our results support recent policy directions such as those stated in the 2022 Public Heath Wales report and the 2021 The Best Start for Life report which highlight that the relationship between the parent and child, between the child’s parents, and the family’s relationships with their wider family, are key components of both children’s wellbeing and early child development [39,40]. Additionally, we add to literature that has found parenting and family support, parenting warmth and parenting styles to be important determinants of psychiatric comorbidity [41]. These results highlight the importance of current UK policy interventions and research endeavours such as ‘A Better Start’, ‘Family Hubs’ and ‘Start for Life’ programmes that aim to give children and families the best start in life [42,43]. Identifying and supporting vulnerable families to develop parenting skills, ensuring children have a healthy and safe home environment are all interventions that may reduce multimorbidity in later life. In addition, continued efforts are needed to address the wider determinants of health such as income and equitable access to good quality housing and healthy food to support people to have healthy lives for longer.

Calculating domain-specific risk scores and using these within the same prediction model was our approach to reduce dimensionality of the individual variables within the domain, while weighing their components, preserving the structure of the key domains, and providing the opportunity to move beyond simple counts of binary variables as an indicator of risk. Additionally, this method was developed building on previous work that derived each variable (within each domain) into a binary (yes/no) outcome, and then summed these binary variables to produce domain risk scores that described a count of adversity [44]. A limitation of this previous approach was that we were required to assume that all variables within each domain and all domain risk scores carried equal weight, and this was particularly problematic when we compared the relative importance of each domain adjusting for the other domains. A further issue was that by deriving variables into a binary indicator we disregarded information contained within the original data structure and we were required to implement arbitrary cut-off points. The methods developed here represent an alternative method that adjusted for the weighting of the individual domain risk scores (in the final prediction model), whilst maintaining the original data structure of the variables.

### Strengths and limitations

Data from a large cohort study allowed us to capture a wide array of biological, social, environmental, behavioural and family variables in childhood to represent five early lifecourse domains. This depth of information would not have been available from most electronic health care records in either primary or secondary care. The data also afforded the opportunity to analyse objective measures of both obesity and hypertension.

However, the cohort is representative of births occurring in Britain in 1970 and as such lacks ethnic diversity. Additionally, no differentiation was made between the burdensome impact of different diseases or disease severity to the individual. For example, mild controlled hypertension or just making the threshold for obesity is unlikely to have the same impact on an individual’s quality of life compared to unmanaged hypertension or clinically severe obesity (BMI over 40). Further, despite using measured rather than self-reported BMI, the BMI measurement continues to have a risk of overestimating body fat in those who have muscular builds.

It is also important to consider the possibility that we have over-adjusted within our models, resulting in overadjustment bias through adjusting for mediators or colliders [45]. This was unavoidable given our particular approach to the analysis given our domains were combined risk factors rather than individual variables. Further, children’s experiences across a variety of early life course domains are intersecting and we argue that research needs to be able to incorporate information from multiple domains into the same analysis. We therefore felt it was important to explore multiple domains simultaneously to attempt to disentangle the strongest predictor for obesity-hypertension comorbidity. We considered taking a more traditional, epidemiological approach where we would consider the relationship between a single domain or a single variable within a domain and odds of obesity-hypertension, controlling for select individual variables from the other domains. However, the choice of domain would have been arbitrary, and this method would not have addressed this paper’s research questions.

Finally, it is important to expand the methods presented here to consider the relationship to other multimorbidity clusters and outcomes that develop a more sophisticated understanding of multimorbidity, including focussing on burdensomeness and complexity. In previous research [27] we audited the early life variables available in two other cohort datasets. The data available in these datasets provide the opportunity to validate and compare our results across cohorts.

### Conclusions

We found the most robust domains for predicting obesity-hypertension were for the parental and family environment domain, the socioeconomic factors domain and the education and academic abilities domain. Developing methods for exploring the multidetermined nature of combined childhood risk factors for health such as the work presented here, can help to challenge existing understanding of the aetiology of health, develop new ideas and solutions, and facilitate improvements in developing and recognising health as a complex and multidetermined concept from a lifecourse perspective.

## Supporting information

Supplementary Docs

## Data Availability

The BCS70 datasets generated and analysed in the current study are available from the UK Data Archive repository (available here: http://www.cls.ioe.ac.uk/page.aspx?&sitesectionid=795).

## Author Contributions

S.F., N.A., R.H., S.P., R.O., S.S. and A.B. contributed to the conceptualisation of the MELD-B project. S.S., N.A., and S.F. obtained the datasets. All authors contributed to the conceptualisation of the paper. S.S., and N.A. led the design and planning of the paper. R.O. led the design of the statistical analysis. S.S., N.A., A.B., N.Z., R.H., and R.O. supported the design, planning and reviewing of the statistical analysis. S.S. performed the statistical analysis with support from N.Z. S.S. prepared all figures and graphs. S.S., and N.A. produced the initial draft of the manuscript. All authors were involved in editing and reviewing the manuscript, and approved the final manuscript. S.S., N.A., and S.F. take responsibility for the data and research governance.

## Acknowledgement

We would like to acknowledge all other members of the MELD-B Consortium, and we thank the participants of the BCS70 cohort study. We would also like to thank Jack Welch and our other PPIE colleagues.

