## Supplementary Docs for "Early Life Domains as Predictors of Obesity and Hypertension Comorbidity: Findings from the 1970 British Cohort Study (BCS70)"

Supplementary Materials

*Supplementary Materials Table 1. All variables that were initially considered for selection within each domain*

Prenatal, antenatal, neonatal and birth domain  (variables recorded at birth)

Maternal age at time of cohort member’s birth

Parity

Labour duration

Number of antenatal visits

Ever a teenage mother

Maternal smoking during pregnancy

Birthweight

Baby illness at birth

Congenital abnormality

Any operations following birth

Method of delivery

Developmental attributes and behaviour domain (variables recorded at age 10)

Number of steps when walking backwards

Balance when standing on right leg

Balance when standing on left leg

Difficulty when kicking a ball

Poor hand control

Clumsy when playing games

Generally clumsy

Does child display outbursts of temper

Does child have emotional or behavioural problems

Rutter behaviour^1^

Difficulty when picking up objects

Socioeconomic factors domain (variables recorded at age 10)

Parental occupational social class^2^

Parental van/car ownership

Cohort member lives on a council estate

Father employment

Mother employment

Income

Housing tenure

Household amenities^3^

Accommodation type

Child education and academic ability domain (variables recorded at age 10)

Edinburgh reading test score

Math ability math – cohort member rating

Reading ability – cohort member rating

Difficult reading – maternal rating

Difficulty writing – maternal rating

Estimated reading age

Friendly math test score

Difficulty with maths – maternal rating

Spelling ability – cohort member rating

Good at spelling – cohort member rating

Parental and family environment domain (variables recorded at age 10)

Family walks

Father plays a role managing the child

Father interested in child education – teacher rating

Mother interested in child education – teacher rating

Family shopping

Father figure

Family goes to restaurants

Parents discuss cohort members education with teacher – teacher rating

Family chat together for 5 mins

Family outings

Family meals

Family holidays

Mother has a dismissive attitude towards cohort member – teacher rating

Father has a dismissive attitude towards cohort member – teacher rating

*^1^Total Score for Rutter behaviour scale*

*^2^Father social class used if available, otherwise mother’s social class used*
*^3^Bathroom/kitchen*

*Supplementary Materials Table 2. Pearson’s Correlation Matrix. Complete case.*

|  | R Child education and academic ability domain predicted probability | Developmental attributes and behaviour domain predicted probability | Prenatal, antenatal, neonatal and birth domain predicted probability | Parental and family environment domain predicted probability | Socio-economic factors domain predicted probability |
| --- | --- | --- | --- | --- | --- |
| Child education and academic ability domain predicted probability | 1.00 |  |  |  |  |
| Developmental attributes and behaviour domain predicted probability | 0.28 | 1.00 |  |  |  |
| Prenatal, antenatal, neonatal and birth domain predicted probability | 0.15 | 0.08 | 1.00 |  |  |
| Parental and family environment domain predicted probability | 0.20 | 0.10 | 0.22 | 1.00 |  |
| Socioeconomic factors domain predicted probability | 0.24 | 0.13 | 0.20 | 0.22 | 1.00 |
| *Sample* | *3955* | | | | |

*Supplementary Materials Figure 1. Step 2: Histogram of the domain predicted risk score (generated in step 2), following logistic regression modelling that explored the relationship between retained variables following stepwise backwards elimination and obesity-hypertension comorbidity for the five early life domains^1^. Multiple imputed data (50 Imputations).*

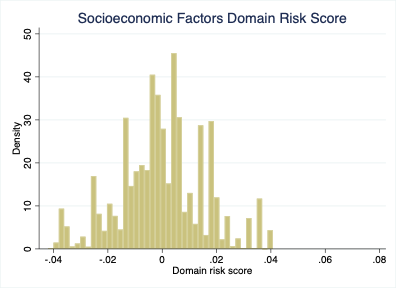

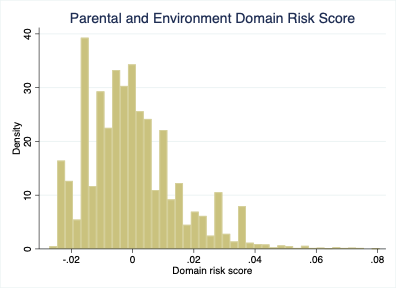

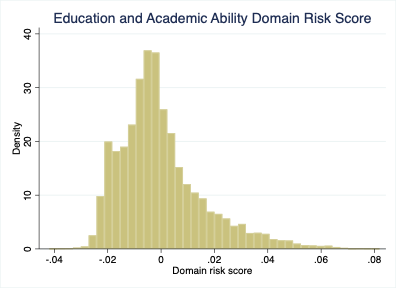

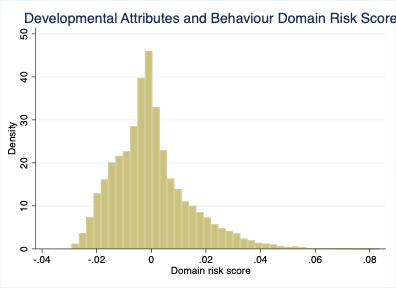

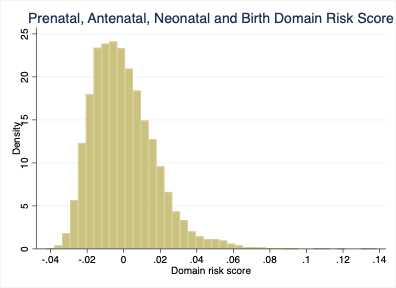

*^1^Domain risk scores were centred on the mean predicted risk score (for each domain) and bound between -1 and 1.*

*Supplementary Materials Table 3. Pearson’s Correlation Matrix. Multiple imputation*

|  | R Child education and academic ability domain predicted probability | Developmental attributes and behaviour domain predicted probability | Prenatal, antenatal, neonatal and birth domain predicted probability | Parental and family environment domain predicted probability | Socio-economic factors domain predicted probability |
| --- | --- | --- | --- | --- | --- |
| Child education and academic ability domain predicted probability | 1.00 |  |  |  |  |
| Developmental attributes and behaviour domain predicted probability | 0.29 | 1.00 |  |  |  |
| Prenatal, antenatal, neonatal and birth domain predicted probability | 0.20 | 0.13 | 1.00 |  |  |
| Parental and family environment domain predicted probability | 0.21 | 0.18 | 0.27 | 1.00 |  |
| Socioeconomic factors domain predicted probability | 0.25 | 0.18 | 0.27 | 0.32 | 1.00 |
| *Sample* | *3955* | | | | |

*Supplementary Materials Table 4. Regression coefficients of the stepwise backward elimination model of the relationship between the antenatal, neonatal and birth domain variables with obesity-hypertension comorbidity. Multiple imputed data (50 Imputations).*

|  | | Coef. | | St.Err. | t-value | p-value | | [95% Conf | Interval] | | Sig |
| --- | --- | --- | --- | --- | --- | --- | --- | --- | --- | --- | --- |
| Mother age at birth of cohort member | | -.018 | | .01 | -1.85 | .065 | | -.037 | .001 | | * |
| Birthweight (Grams) | | 0 | | 0 | -2.57 | .01 | | 0 | 0 | | ** |
| Mothers Parity : base 0 | | 0 | | . | . | . | | . | . | |  |
| 1 | | -.101 | | .105 | -0.96 | .335 | | -.306 | .104 | |  |
| 2 | | 0 | | .138 | 0.00 | .997 | | -.271 | .271 | |  |
| 3 | | -.038 | | .201 | -0.19 | .85 | | -.432 | .356 | |  |
| 4 | | .476 | | .243 | 1.96 | .05 | | -.001 | .953 | | * |
| 5+ | | .499 | | .288 | 1.73 | .084 | | -.067 | 1.064 | | * |
| Maternal smoking during pregnancy: base non smoker | | 0 | | . | . | . | | . | . | |  |
| Stopped Pre/During pregnancy | | .078 | | .125 | 0.62 | .533 | | -.167 | .323 | |  |
| Smoker | | .234 | | .096 | 2.45 | .014 | | .047 | .422 | | ** |
| Constant | | -1.444 | | .362 | -3.98 | 0 | | -2.154 | -.733 | | *** |
| Imputations | 50 | | Number of obs | | | | 7858 | | |  |  |
| Prob > F | 0.0010 | |  | | | |  | | |  |  |
| F( 9, 9. 97e+07) | 3.11 | |  | | | |  | | |  |  |
| **** p<.01, ** p<.05, * p<.1* | | | | | | | | | | | |

*.*

*Supplementary Materials Table 5. Regression coefficients of the stepwise backward elimination model of the relationship between the developmental attributed and behaviour domain variables with obesity-hypertension comorbidity. Multiple imputed data (50 Imputations).*

|  | | Coef. | | St.Err. | t-value | | p-value | [95% Conf | | Interval] | Sig |
| --- | --- | --- | --- | --- | --- | --- | --- | --- | --- | --- | --- |
| Rutter Behavior | | 0 | | 0 | 2.20 | | .028 | 0 | | .001 | ** |
| Number of steps walking backwards | | -.019 | | .008 | -2.36 | | .018 | -.035 | | -.003 | ** |
| Hand control rating | | -.012 | | .004 | -3.16 | | .002 | -.019 | | -.004 | *** |
| Constant | | -2.069 | | .195 | -10.63 | | 0 | -2.45 | | -1.687 | *** |
| Imputations | 50 | | Number of obs | | | 7858 | | |  |  |  |
| Prob > F | 0.0000 | |  | | |  | | |  |  |  |
| F( 3, 6710.2) | 7.93 | |  | | |  | | |  |  |  |
| **** p<.01, ** p<.05, * p<.1* | | | | | | | | | | | |

*Supplementary Materials Table 6. Regression coefficients of the stepwise backward elimination model of the relationship between the socioeconomic factor domain variables with obesity-hypertension comorbidity. Multiple imputed data (50 Imputations).*

|  | Coef. | | St.Err. | t-value | | p-value | [95% Conf | | Interval] | | Sig |
| --- | --- | --- | --- | --- | --- | --- | --- | --- | --- | --- | --- |
| Parental social class: base I professional | 0 | | . | . | | . | . | | . | |  |
| II managerial | -.045 | | .228 | -0.20 | | .842 | -.492 | | .402 | |  |
| III non manual | .085 | | .256 | 0.33 | | .74 | -.416 | | .586 | |  |
| III manual | .198 | | .232 | 0.85 | | .394 | -.257 | | .652 | |  |
| IV partly-skilled | .405 | | .249 | 1.63 | | .104 | -.084 | | .894 | |  |
| V unskilled | .109 | | .339 | 0.32 | | .746 | -.555 | | .774 | |  |
| Housing tenure: base Owned outright | 0 | | . | . | | . | . | | . | |  |
| Rented, private | -.075 | | .31 | -0.24 | | .808 | -.683 | | .532 | |  |
| Being purchased | .07 | | .154 | 0.46 | | .647 | -.231 | | .372 | |  |
| Rented, Council | .201 | | .165 | 1.22 | | .222 | -.122 | | .524 | |  |
| Other | .111 | | .29 | 0.38 | | .701 | -.458 | | .681 | |  |
| Gross income per week: base £250+ | 0 | | . | . | | . | . | | . | |  |
| £200 - £249 | .524 | | .301 | 1.74 | | .082 | -.066 | | 1.113 | | * |
| £150 - £199 | .244 | | .275 | 0.89 | | .375 | -.295 | | .784 | |  |
| £100 - £49 | .49 | | .261 | 1.88 | | .061 | -.023 | | 1.003 | | * |
| £50 - £99 | .529 | | .263 | 2.01 | | .044 | .013 | | 1.044 | | ** |
| under £49 | .571 | | .313 | 1.82 | | .068 | -.043 | | 1.184 | | * |
| Constant | -3.189 | | .318 | -10.03 | | 0 | -3.812 | | -2.565 | | *** |
| Imputations | | 50 | | | Number of obs | | | 7858 | |  |  |
| Prob > F | | 0.0062 | | |  | | |  | |  |  |
| F( 14, 45629.9) | | 2.19 | | |  | | |  | |  |  |
| **** p<.01, ** p<.05, * p<.1* | | | | | | | | | | | |

*Supplementary Materials Table 7. Regression coefficients of the stepwise backward elimination model of the relationship between the education and academic ability domain variables with obesity-hypertension comorbidity. Multiple imputed data (50 Imputations).*

|  | | Coef. | | St.Err. | t-value | p-value | | [95% Conf | Interval] | | Sig |
| --- | --- | --- | --- | --- | --- | --- | --- | --- | --- | --- | --- |
| Edinburgh Reading Test Scores | | -.012 | | .004 | -2.89 | .004 | | -.02 | -.004 | | *** |
| Difficulty reading – teacher’s assessment: base no difficulty | | 0 | | . | . | . | | . | . | |  |
| Some difficulty | | .097 | | .132 | 0.74 | .461 | | -.162 | .357 | |  |
| Great difficulty | | -.322 | | .344 | -0.94 | .349 | | -.997 | .353 | |  |
| Ability spelling – cohort members assessment: base good ability | | 0 | | . | . | . | | . | . | |  |
| Not so well | | .185 | | .097 | 1.90 | .058 | | -.006 | .375 | | * |
| Difficulty writing – teacher’s assessment: base no difficulty | | 0 | | . | . | . | | . | . | |  |
| Some difficulty | | .211 | | .12 | 1.76 | .079 | | -.024 | .446 | | * |
| Great difficulty | | -.247 | | .418 | -0.59 | .556 | | -1.068 | .575 | |  |
| Constant | | -2.134 | | .2 | -10.66 | 0 | | -2.527 | -1.741 | | *** |
| Imputations | 50 | | Number of obs | | | | 7858 | | |  |  |
| Prob > F | 0.0001 | |  | | | |  | | |  |  |
| F( 6, 12508.3) | 4.87 | |  | | | |  | | |  |  |
| **** p<.01, ** p<.05, * p<.1* | | | | | | | | | | | |

*Supplementary Materials Table 8. Regression coefficients of the stepwise backward elimination model of the relationship between parental and family environment domain variables with obesity-hypertension comorbidity. Multiple imputed data (50 Imputations).*

|  | Coef. | | St.Err. | t-value | | p-value | [95% Conf | | Interval] | | Sig |
| --- | --- | --- | --- | --- | --- | --- | --- | --- | --- | --- | --- |
| Father Figure: base biological father | 0 | | . | . | | . | . | | . | |  |
| Father by adoption | .377 | | .344 | 1.09 | | .274 | -.298 | | 1.052 | |  |
| Step father | .282 | | .216 | 1.31 | | .19 | -.14 | | .705 | |  |
| Other | .319 | | .324 | 0.98 | | .326 | -.317 | | .955 | |  |
| No father figure | -.021 | | .311 | -0.07 | | .947 | -.631 | | .59 | |  |
| Family go on outings: base rarely or never | 0 | | . | . | | . | . | | . | |  |
| Sometimes | .151 | | .259 | 0.58 | | .56 | -.357 | | .66 | |  |
| Often | .066 | | .261 | 0.25 | | .801 | -.445 | | .577 | |  |
| Father helps to manage cohort member base: equal or similar amount | 0 | | . | . | | . | . | | . | |  |
| Smaller part than that mother | -.168 | | .103 | -1.62 | | .105 | -.37 | | .035 | |  |
| Very small part | -.137 | | .184 | -0.74 | | .457 | -.497 | | .223 | |  |
| Does not help | .024 | | .295 | 0.08 | | .935 | -.554 | | .602 | |  |
| Mother’s interest in education: base very interested | 0 | | . | . | | . | . | | . | |  |
| Moderate Interest | .082 | | .137 | 0.60 | | .549 | -.187 | | .351 | |  |
| Very Little Interest | .231 | | .264 | 0.87 | | .382 | -.287 | | .748 | |  |
| Uninterested | .336 | | .438 | 0.77 | | .443 | -.524 | | 1.197 | |  |
| Cannot say/no mother | .293 | | .171 | 1.72 | | .087 | -.042 | | .628 | | * |
| Father’s interest in education: base very interested | 0 | | . | . | | . | . | | . | |  |
| Moderate Interest | .136 | | .157 | 0.87 | | .386 | -.173 | | .446 | |  |
| Very Little Interest | .208 | | .298 | 0.70 | | .485 | -.377 | | .793 | |  |
| Uninterested | -.026 | | .393 | -0.07 | | .948 | -.799 | | .747 | |  |
| Cannot say/no father | .274 | | .146 | 1.87 | | .062 | -.014 | | .561 | | * |
| Constant | -2.778 | | .269 | -10.34 | | 0 | -3.305 | | -2.251 | | *** |
| Imputations | | 50 | | | Number of obs | | | 7858 | |  |  |
| Prob > F | | 0.0742 | | |  | | |  | |  |  |
| F( 17, 25109.5) | | 1.53 | | |  | | |  | |  |  |
| **** p<.01, ** p<.05, * p<.1* | | | | | | | | | | | |

*Supplementary Materials Table 9. Odds of obesity-hypertension at age 46 in relation to predicted probabilities of obesity-hypertension for five early life domains. Complete Case.*

|  | Model adjusting for confounders | | Model adjusting for confounders and other domains | |
| --- | --- | --- | --- | --- |
|  | OR | 95%CI | OR | 95%CI |
| Child education and academic ability domain predicted probability | **1.11** | **1.06-1.16** | 1.03 | 0.98-1.09 |
| Developmental attributes and behaviour domain predicted probability | **1.11** | **1.06-1.16** | **1.08** | **1.03-1.13** |
| Prenatal, antenatal, neonatal and birth domain predicted probability | **1.14** | **1.07-1.20** | **1.07** | **1.01-1.13** |
| Parental and family environment domain predicted probability | **1.12** | **1.07-1.17** | **1.08** | **1.04-1.12** |
| Socioeconomic factors domain predicted probability | **1.14** | **1.08-1.20** | **1.08** | **1.03-1.14** |
| *Sample* | *3965* | | | |

*Supplementary Materials 10. Receiver operating characteristic curve for prediction model including all five domain specific risk scores, sex and ethnicity in the same model. Multiple imputed data (50 Imputations).*

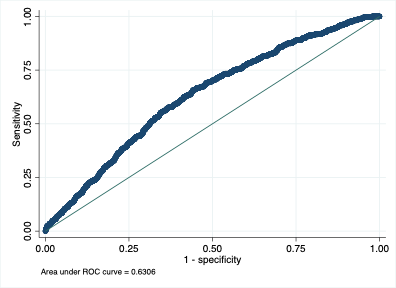

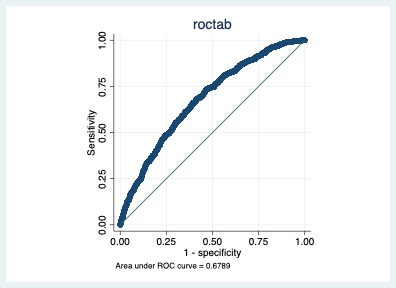
*Supplementary Materials 11. Receiver operating characteristic curve for prediction model including all five domain specific risk scores, sex and ethnicity in the same model. Multiple imputed data (50 Imputations).*
